# Relationships between the auditory nerve’s ability to recover from neural adaptation, cortical encoding of and perceptual sensitivity to within-channel temporal gaps in postlingually deafened adult cochlear implant users

**DOI:** 10.1101/2022.10.20.22281267

**Authors:** Shuman He, Yi Yuan, Jeffrey Skidmore

## Abstract

**Objective:** This study assessed the relationships between the auditory nerve’s ability to recovery from neural adaptation, cortical encoding of and perceptual sensitivity to within-channel temporal gaps in postlingually deafened adult cochlear implant (CI) users.

**Design:** Study participants included 11 postlingually deafened adults with Cochlear™ Nucleus® devices, including three bilaterally implanted participants. In each participant, recovery from neural adaptation of the auditory nerve (AN) was measured using electrophysiological measures of the electrically evoked compound action potential (eCAP) at up to four electrode locations. The electrode pair with the largest difference in the speed of adaptation recovery was selected for assessing within-channel temporal gap detection threshold (GDT). GDTs were measured using both psychophysical and electrophysiological procedures. Psychophysical GDTs were evaluated using a three-alternative, forced-choice procedure, targeting 79.4% correct on the psychometric function. Electrophysiological GDTs were measured using the electrically evoked auditory change complex (eACC) evoked by electrical pulse trains that contained temporal gaps. Objective GDT was defined as the shortest temporal gap that could evoke an eACC. Related-samples Wilcoxon Signed Rank testwas used to compare psychophysical GDTs and objective GDTs measured at all electrode locations. It was also used to compare psychophysical GDTs and objective GDTs measured at the two electrode locations with different speeds or amounts of adaptation recovery of the AN. A Kendall Rank correlation test was used to assess the correlation between GDTs measured using psychophysical or electrophysiological procedures.

**Results:** Objective GDTs were significantly larger than those measured using psychophysical procedures. There was a significant correlation between objective and psychophysical GDTs. GDTs could not be predicted based on the amount or the speed of adaptation recovery of the AN.

**Conclusions:** Electrophysiological measure of the eACC can potentially be used to assess within-channel GDT in CI users who cannot provide reliable behavioral responses. The difference in adaptation recovery of the AN is not the primary factor accounting for the across-electrode variation in GDT in individual CI users.

## INTRODUCTION

Auditory temporal resolution refers to the ability of the auditory system to follow rapid changes in auditory inputs over time. It plays a crucial role in speech perception, especially in patients with cochlear implants (CIs) due to the limited number of independent spectral channels provided by the CI (e.g., Shannon et al., 1995; Fu et al., 2004). In CI users, higher temporal resolution sensitivities are associated with better speech perception performance (e.g., Fu, 2002; Luo et al., 2008, 2020; He et al., 2013; Erb et al., 2019). The gap detection threshold (GDT) has been used to assess temporal resolution sensitivity in CI users in many studies (e.g., Wei et al., 2007; He et al., 2013, 2018a; Mussoi & Brown, 2019; Luo et al., 2020; Tuz et al., 2020). It refers to the shortest silence (temporal gap) embedded in auditory signals that can be detected by human listeners. The GDT can be evaluated using psychophysical procedures (e.g., Wei et al., 2007; Luo et al., 2020; Tuz et al., 2020), as well as electrophysiological measures of the electrically evoked auditory change complex (eACC) (e.g., He et al., 2013, 2018a; Mussoi & Brown, 2019). The eACC is an obligatory event-related potential (eERP) that is generated in the central auditory system of implanted patients. It can be elicited by stimulus change(s) occurring within an ongoing, long-duration electrical stimulation. The presence of the eACC provides an indication of auditory discrimination capability at the auditory cortex (Martin et al., 2008). In adult CI users, the eACC consists of a P1-N1-P2 complex, which the same as the eERP evoked by a stimulus onset (i.e., the onset eERP, e.g., Brown et al., 2015; Mussoi & Brown, 2019; van Heteren et al., 2022).

In CI users, GDTs measured using both psychophysical and electrophysiological procedures show substantial inter-patient variations, as well as across-electrode variations within individual patients (e.g., Pfingst et al., 2008; Garadat & Pfingst. 2011; Bierer et al., 2015; He et al., 2018a). These variations have been attributed to the differences in local cochlea/auditory nerve (AN) health near stimulation sites (Pfingst et al., 2008; Bierer et al., 2015). In human CI users, it is not feasible to use histological measures to directly assess cochlea/AN health. However, useful information about the functional status (the number and the responsiveness of surviving neural element) of the AN can be derived from the results of electrophysiological measures of the electrically evoked compound action potential (eCAP) (e.g., Long et al., 2014; He et al., 2018b, 2022a, 2022b; Shader et al., 2020; Skidmore et al., 2021). The eCAP represents the synchronized response of a population of electrically stimulated AN fibers. eCAP parameters that have been associated with the functional status of the AN include slope of the eCAP input/output (I/O) function (e.g., Miller et al., 1994; Pfingst et al., 2015a, 2015b; Pfingst et al., 2017), sensitivity of the eCAP to changes in interphase gap (e.g., Prado-Guitierrez et al., 2006; Ramekers et al., 2014; Schvartz-Leyzac & Pfingst, 2018; Schvartz-Leyzac et al., 2019; He et al., 2019; Skidmore & He, 2021) and phase polarity (Macherey et al., 2008; Undurraga et al., 2010) of a biphasic pulse, as well as the speed of recovery from refractoriness (He et al., 2018b) and neural adaptation (Ramekers et al., 2015; He et al., 2022b).

To date, there has only been one study that measured eCAPs and GDTs at the same group of stimulation sites in CI users. Specifically, Shader et al. (2020) measured slopes of the eCAP I/O function and GDTs at five CI electrode locations in thirty adult CI users. Their results showed that sleeper slopes were significantly correlated with smaller GDTs in adult CI users who were 45 years of age or younger when results measured at different pulse rates and electrode locations were averaged together. However, this significant relationship was not observed in middle-age (46-64 years of age) or older (65-80 years of age) CI users. More importantly, there was no direct assessment of the relationship between GDT and slope of the eCAP I/O function measured at the same electrode location within each participant. Consequently, results of that study did not provide conclusive evidence supporting the direct link between local cochlea/AN health and GDT result.

The slope of the eCAP I/O function is measured using single-pulse stimulation. In comparison, pulse-train stimulation is typically used to measure psychophysical and electrophysiological GDTs. It has been shown that using single pulses for electrophysiological recordings and pulse trains for psychophysical assessments confounds the potential relationship between results measured using these two procedures (Adel et al., 2017). Therefore, eCAP measures using pulse-train stimulation, such as those used to assess neural adaptation and recovery from neural adaptation (for a review see He et al., 2017), may be a better choice than eCAPs evoked by single-pulses for investigating the relationship between local AN health and GDT results. This idea is based on the important roles of these two temporal response properties of the AN in neural encoding of temporal envelope cues, including temporal gaps. Specifically, temporal cues, especially rapid amplitude changes, are represented in the discharge patterns of the AN (Delgutte, 1980; Delgutte & Kiang, 1984). The firing rate of the AN rapidly increases to the maximum at the onset of sustained stimulation followed by a gradual decay in instantaneous firing rate (i.e., neural adaptation). Due to neural adaptation, responsiveness to subsequent stimulation is reduced for a brief period following the cessation of the initial stimulation, resulting in forward masking effects. Normal neural adaptation and adaptation recovery have been proposed to have the important roles of producing peaks in the discharge rate of the AN that serve to enhance acoustic onsets in the speech waveform (Delgutte, 1997). Aberrant neural adaptation and adaptation recovery can lead to smeared representation of temporal envelope cues at the AN, which could lead to prolonged GDTs and poor speech perception performance. In alignment with this theoretical possibility, slow adaptation recovery has recently been shown to be associated with poor speech perception in CI users (He et al., 2022b).

The association between neural adaptation of the AN and psychophysical GDTs has been investigated in 14 adult CI users by Zhang et al. (2013). Their results showed a lack of significant correlation between the amount of neural adaptation of the AN and GDT. Neural adaptation recovery of the AN in adult CI users has been recently characterized by He et al. (2022b). Similar with GDT results reported in the literature, the speed and the amount of adaptation recovery also showed substantial variations across different CI electrode sites within individual CI users. To date, the association between adaptation recovery of the AN and GDT has not been investigated in CI users. As a result, their relationship remains unknown.

In summary, GDTs measured in CI users demonstrate substantial variations among patients, as well as variations across electrode locations within individual patients. The neurophysiological mechanism underlying these variations remains unknown. To address this critical knowledge gap, this study investigated the relationship between neural adaptation recovery of the AN and psychophysically and electrophysiologically measured GDT in postlingually deafened adult CI users. It tested the hypothesis that the difference in neural adaptation recovery of the AN accounted for the variations in GDTs measured at different stimulation sites within individual CI users. It was expected that less recovery and/or prolonged recovery from neural adaptation of the AN would lead to lower temporal resolution sensitivities, as evidenced by higher GDTs, in human CI users.

## MATERIALS AND METHODS

### Study Participants

Study participants included three bilaterally implanted and eight unilaterally implanted postlingually deafened adult CI users, including four males and seven females. All 14 implanted ears were tested in this study. All participants used a Cochlear™ Nucleus® device (Cochlear Ltd., Macquarie, NSW, Australia) with full electrode insertion in the test ear(s), were native speakers of American English, and scored 26 or higher on the Mini-Mental State Examination (Folstein et al., 1975). Age at implantation of the test ear ranged from 25.6 to 77.71 years (mean: 56.98 yrs, SD: 15.17 yrs). Each participant was tested for multiple sessions. Age at initial testing ranged from 36.79 to 79.39 years (mean: 62.85 yrs, SD: 13.54 yrs).

Detailed demographic information of these participants is listed in Table 1. The study was approved by the local Biomedical Institutional Review Board (2017H0131). Written, informed consents were obtained from study participants prior to participation. All participants received monetary compensation for participating in this study.

**Table 1.** Demographic information of study participants. L: left, R: right, AAI: age at implantation, AAT: age at testing, 24RE (CA): Freedom Contour Advance electrode array, eCAP: electrically evoked compound action potential, AR: adaptation recovery, GDT: gap detection threshold.

| **Subject number** | **Ear tested** | **AAI (yrs)** | **AAT (yrs)** | **Internal electrode array** | **Electrodes with artifact-free eCAPs tested for AR** | **Electrode tested for G** |
| --- | --- | --- | --- | --- | --- | --- |
| A1 | L | 50s | 60s | CI512 | 4, 9 and 15 | 4 and 15 |
| A2 | L | 50s | 60s | 24RE (CA) | 3, 9, 15 and 21 | 3 and 21 |
| A2 | R | 50s | 60s | 24RE (CA) | 3, 9, 15 and 21 | 9 and 21 |
| A3 | L | 30s | 30s | 24RE (CA) | 3, 9, 15 and 21 | 9 and 21 |
| A4 | L | 50s | 50s | CI532 | 4, 9, 15 and 21 | 15 and 21 |
| A4 | R | 40s | 50s | 24RE (CA) | 3, 9, 15 and 21 | 9 and 21 |
| A5 | R | 60s | 60s | CI522 | 4, 8, 15 and 21 | 4 and 21 |
| A6 | R | 20s | 30s | 24RE (CA) | 3, 9, 15 and 21 | 3 and 21 |
| A7 | R | 60s | 60s | CI532 | 15 and 21 | 15 and 21 |
| A8 | L | 70s | 70s | CI422 | 4, 10, 15 and 20 | 4 and 20 |
| A9 | L | 60s | 70s | 24RE (CA) | 7, 16, 18 and 21 | 7 and 18 |
| A10 | L | 70s | 70s | CI622 | 3, 9, 15 and 21 | 15 and 21 |
| A10 | R | 70s | 70s | CI522 | 4, 9, 15 and 20 | 15 and 20 |
| A11 | L | 50s | 50s | CI532 | 9, 15 and 21 | 9 and 21 |

### Stimulus

The stimulus was a train of biphasic pulses with a carrier rate of 900 pulses per second (pps) per channel. The pulse phase duration and the interphase gap of each biphasic pulse was 25 μs and 7 μs, respectively. In this study, stimuli used in all measures were directly delivered to individual CI electrodes via an N6 sound processor interfaced with a programming pod. All stimuli were presented in a monopolar-coupled stimulation mode (MP1) at the maximum comfortable level that was measured for each CI electrode tested in each participant. The stimulus used for eCAP measures was generated using the Custom Sound EP (v.5.1 or v.5.2) commercial software (Cochlear Ltd, Macquarie, NSW, Australia). The stimuli used in psychophysical measures of GDTs and electrophysiological measures of eERPs were created using custom-designed software incorporating Nucleus Interface Communication programming routines (NIC v.4).

## Experimental Procedures

### Overview

For each implanted ear, adaptation recovery of the AN was evaluated using eCAP measures at up to four CI electrodes (typically defaulted to electrodes 3, 9, 15 and 21) across the electrode array. In all except for one (reported below) test ears, psychophysical measures of GDT and electrophysiological measures of the eACC evoked by temporal gaps were obtained at two electrode locations with the largest difference in the speed of adaptation recovery of the AN. The speed of adaptation recovery of the AN was used to determine the electrode locations for GDT assessment due to its close association with speech perception performance in CI users (He et al., 2022b). Artifact-free eCAPs were only recorded at two electrode locations in the right ear of A7. As a result, psychophysical GDTs and eACCs were measured at these two electrode locations. CI electrodes tested for eCAP and GDT measures in each participant are listed in Table 1. The trains of biphasic pulses presented before and after temporal gaps were defined using identical parameters. Therefore, GDTs measured in this study are within-channel GDTs.

### eCAP Measures

eCAP recordings were obtained using the Advanced Neural Response Telemetry (NRT) function implemented in the Custom Sound EP (v.5.1 or v.5.2) commercial software (Cochlear Ltd, Macquarie, NSW, Australia). eCAPs were recorded with a 122-μs recording delay, a 20-kHz sampling rate, an amplifier gain of 50 dB and 50 sweeps per averaged response.

Details of using the eCAP to assess neural adaptation recovery of the AN have been reported in He et al. (2022a). Briefly, eCAPs evoked by the probe pulse presented at 1.054, 2, 4, 8, 16, 32, 64, 128 and 256 ms after the offset of the last pulse of the pulse-train (i.e., the masker) were recorded using a modified forward-masking technique (He et al., 2017). As the time interval between the masker and the probe (i.e., masker-probe-interval, MPI) increased, AN fibers gradually recovered from the neural adaptation induced by the pulse-train masker, which resulted in gradually increased eCAP amplitudes at longer MPIs.

### Psychophysical Measures of Gap Detection Threshold

The stimuli were 500-ms biphasic pulse trains with and without temporal gaps. They were presented in a three-interval, two-alternative, forced-choice paradigm that incorporated a three-down, one-up adaptative strategy estimating 79.4% correct detection (Levitt, 1971). The duration of each listening interval and the interstimulus interval was 500 ms. The stimulus presented in two listening intervals was a 500 ms pulse train without any interruption. The stimulus presented in the third interval, chosen at random, included a temporal gap centered at 250 ms of stimulation. The participant was asked to determine which interval included two sounds. No feedback was given for incorrect responses. The initial step size of the change in gap duration was 32 ms. This step changed by a factor of two after three consecutive correct responses or one incorrect response until the gap duration was 1 ms or 256 ms. A threshold track stopped after 12 reversals, and the gap duration at the final four reversals was geometrically averaged. For each CI electrode tested in each participant, two estimates were obtained, and behavioral GDT was defined as the average of these two estimates.

### eERP Measures

The stimulus was an 800-ms biphasic pulse train presented at 0.5 Hz in two stimulation conditions: the *standard* condition and the *gapped* condition. In the *standard* condition, the 800-ms pulse train was presented to the selected CI electrode without any interruption. In the *gapped* condition, a temporal gap was inserted after 400 ms of stimulation. Gap durations tested in this study ranged from 1 up to 64 ms in logarithmic steps.

Electroencephalographic (EEG) activity was recorded using a Brain Vision system (BrainAmp DC) and disposable, sterile Ag-AgCI surface recording electrodes. The EEG was recorded differentially between five active recording electrodes (Fz, FCz, Cz, C3 and C4) and contralateral mastoid (A^1/2^, reference) relative to a ground electrode placed at the low forehead (Fpz). Electrode impedances were below 5 kΩ and the inter-electrode impedance difference was less than 2 kΩ. The recording window was 1500 ms, including a 100-ms pre-stimulus baseline. The EEG was sampled at 1000 Hz, baseline corrected, and online filtered between 0.1 and 100 Hz (12 dB/octave roll off). Eye movements were monitored using a pair of recording electrodes placed above and below the eye that was contralateral to the CI. Responses exceeding 100 μV were rejected from averaging. For each participant, at least three replications with 100 artifact-free sweeps per replication were recorded for each stimulating condition. These replications were digitally filtered between 1 – 30 Hz (12 dB/octave) offline. Replications recorded for the same stimulating condition were averaged together. Individual replications along with the averaged response were used to determine the presence/absence of the eACC.

All participants were tested while seated in a comfortable chair and watching a silent movie with captions. Participants were instructed to be relaxed and stay awake. Breaks were provided as necessary to ensure compliance with these instructions.

### Data Analysis

Approaches used to analyze eCAP results and estimate the speed of adaptation recovery (τ_3_) have been reported in detail in He et al. (2022a). Briefly, artifact-free eCAP results were derived using custom-designed MATLAB (v.2021a, The Mathworks Inc., United States) software. The adaptation recovery function (ARF) was obtained by plotting normalized eCAP amplitudes (re. the eCAP amplitude measured at the MPI of 256 ms) as a function of MPIs on a logarithmic scale. The time constant for neural adaptation recovery (i.e., τ_3_) was estimated using a mathematical model with an exponential function in the form of

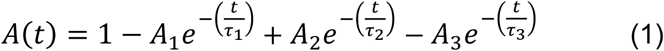

Detailed definitions and descriptions of three types of ARFs recorded in human CI users, as well as different parameters included in this function have been reported in He et al. (2022a). The amount of adaptation recovery was quantified using the adaptation recovery ratio (ARR). It was defined as the ratio between the eCAP amplitude measured at the MPI of 256 ms and the eCAP amplitude evoked by a single pulse presented at the same maximum comfortable level as that of the masker pulse train. Larger ARRs indicated more/greater recovery from neural adaptation.

For eERP results, replicated responses and the averaged response of these replications measured for each stimulation condition at each electrode location were overlapped to show their repeatability. Neural responses were considered present only if all replications recorded for the same stimulation condition at all recording sites were repeatable. Responses were independently evaluated by two judges who were blind to participant’s identification and stimulation condition to determine the presence/absence of the onset eERP and the eACC. The disagreements over the presence/absence of these responses were resolved through discussion. For each participant, the objective GDT was defined as the shortest gap duration that could evoke the eACC. The eACC could not be identified for any gap durations tested at electrode 9 in the left ear of A3. In this case, the objective GDT was assigned to be 65 ms.

The results of Shapiro-Wilk test showed that the distribution of GDTs measured using both psychophysical and electrophysiological procedures departed significantly from normality (psychophysical GDTs: W = 0.722, p < .001; objective GDTs: W = 0.551, p < .001). Therefore, related-samples Wilcoxon Signed rank test was used 1) to compare psychophysical GDTs and objective GDTs, and 2) to compare psychophysical and objective GDTs measured at the electrode locations with smaller τ_3_s or ARRs with those measured at the electrode locations with larger τ_3_s or ARRs. A Kendall Rank correlation test was used to assess the correlation between psychophysical and objective GDTs. All statistical tests were conducted using SPSS Statistic 28 (IBM Corp.).

## RESULTS

Figure 1 shows group means and standard deviations of eERPs recorded at all CI electrode locations for all stimulation conditions. The gap duration tested in each stimulation condition is labeled for these traces. These data clearly show that eERPs recorded in this study are free of artifact contamination. The onset eERP can be identified for each averaged response within the time window between 15 and 330 ms after stimulus onset, as indicated by the rectangle. The eACC can be identified for the averaged responses recorded at gap durations of 8 ms and longer. Both the onset eERP and the eACC included in these traces consist of a P1-N1-P2 complex. The time window of the eACC, as indicated by the parallelogram, varies with gap durations with delayed time windows at longer gap durations. It should be pointed out that the time window of eACCs indicated in Figure 1 was determined based on the results observed in all participants instead of the group averaged data. Therefore, it extends to traces recorded in the 1-ms gap condition.

**Figure 1.**
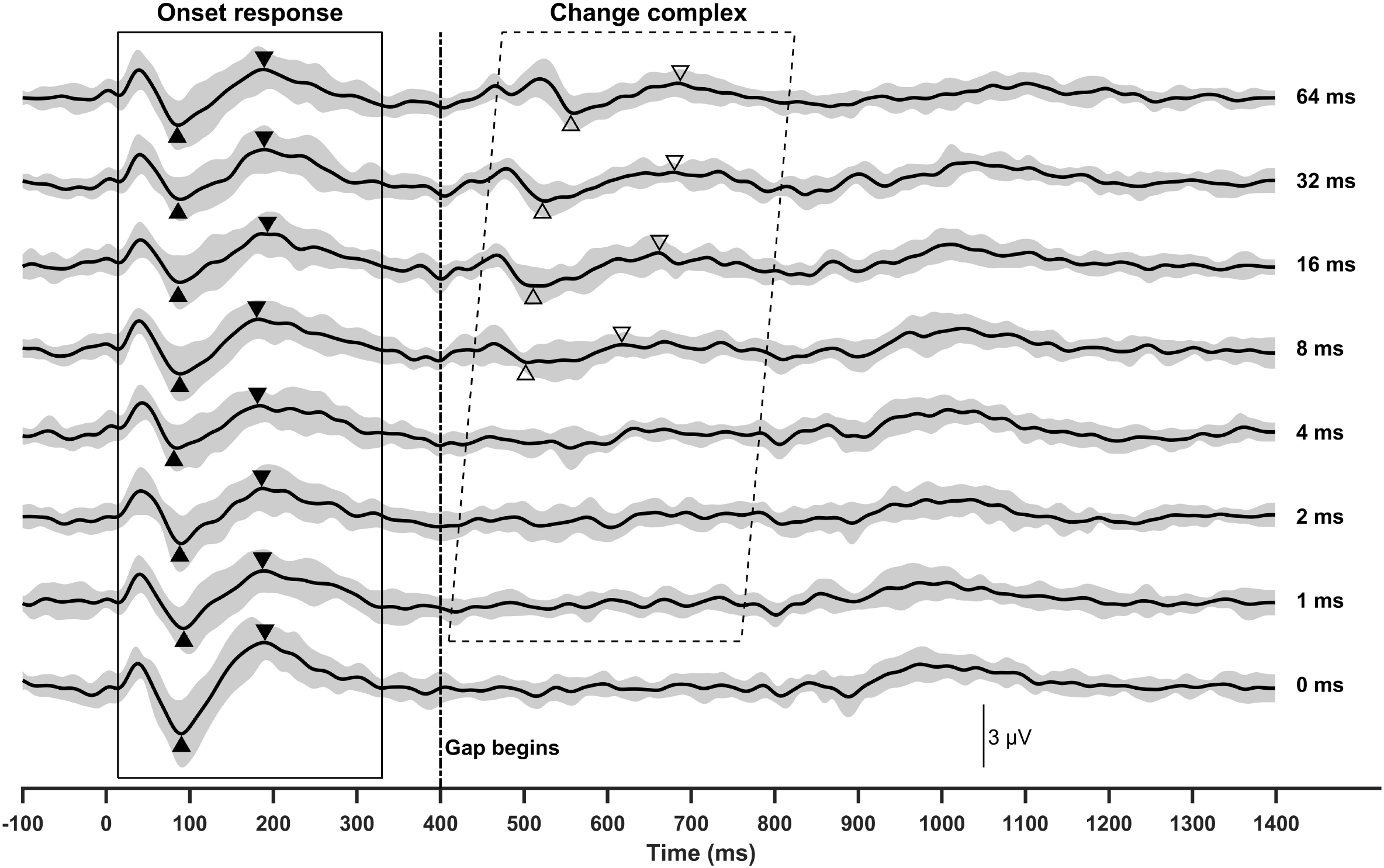
Group averaged eERPs measured for all stimulation conditions at all electrode locations tested in all participants. Shaded areas indicate standard deviations. Filled and open triangles indicate N1 and P2 peaks of the onset eERP and the eACC, respectively. The vertical dashed line indicates the time point when the first 400 ms of stimulation ended.

### GDTs measured using different procedures

GDTs measured using psychophysical procedures ranged from 1.5 to 11.67 ms (mean: 3.30 ms, SD: 2.51 ms). In comparison, GDTs measured using electrophysiological procedures showed a much larger range (1 - 65 ms) with a higher mean value (10.71 ms) and more variations (SD: 16.61 ms). The result of a related-samples Wilcoxon Signed rank test revealed a statistically significant difference between behavioral and objective GDTs (Z = 3.559, p < .001).

Panel (a) of Figure 2 shows objective GDTs plotted as a function of psychophysical GDTs measured in this study. The result of the Kendall Rank correlation test, as indicated in this panel, revealed a statistically significant correlation between GDTs measured using these two procedures. These data clearly showed that objective GDTs measured at three electrode locations were substantially larger than those measured using psychophysical procedures. Specifically, objective GDTs measured at electrodes 9 and 21 in the left ear of A3 were 65 and 32 ms respectively. The objective GDT measured at electrode 4 in the right ear of A5 was 64 ms. In comparison, the corresponding psychophysical GDTs measured at these electrode locations were 11.67, 5.83 and 2.42 ms. The difference between objective and psychophysical GDTs ranged from 26.17 ms to 61.58 ms, which is much larger than the absolute difference between these two values measured at other electrode locations (range: 0 - 13.67 ms, mean: 3.00 ms, SD: 3.06 ms). As a result, these data points might have biased the statistical result. Panel (b) of Figure 2 shows GDT results without these three data points. Based on the result of the Kendall Rank correlation test for this reduced dataset, GDTs measured using psychophysical and electrophysiological procedures remain significantly correlated.

**Figure 2.**
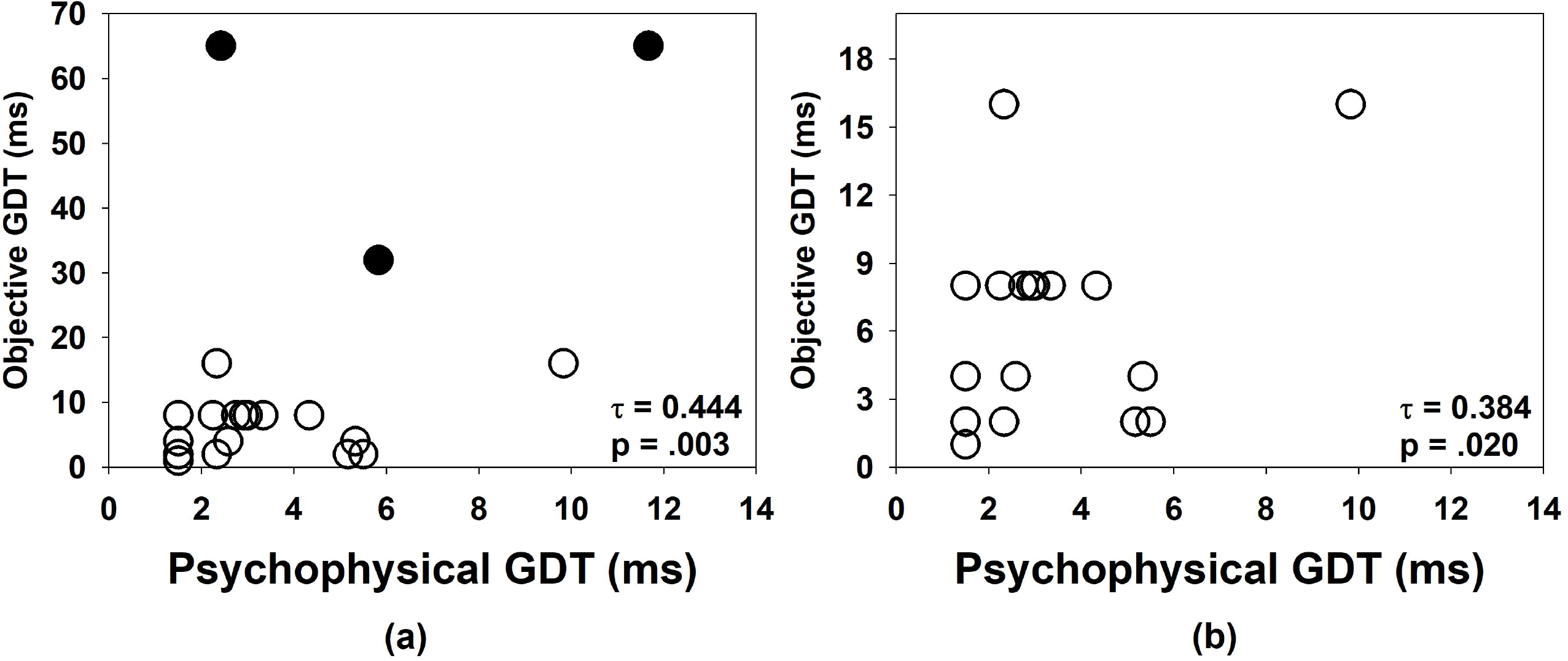
The relationship between GDTs measured using psychophysical and electrophysiological procedures. Each symbol indicates the result measured at one electrode location. Filled circles in panel (a) indicate the results measured in three ears that showed much higher objective GDTs than psychophysical GDTs.

Figure 3 depicts eERPs recorded at the three CI electrode locations in two participants who showed substantially larger objective GDTs than those measured using psychophysical procedures. The onset eERP could be easily identified for all averaged responses. In addition, a total of 1200 sweeps were recorded for each stimulation condition tested at electrode 4 in A5. Despite such a large number of accepted sweeps, the eACC was not identified for gap durations of 32 ms or shorter. Overall, these results suggested that the lack of the eACC response at some gap conditions was not due to inadequate stimulation level or insufficient number of accepted sweeps.

**Figure 3.**
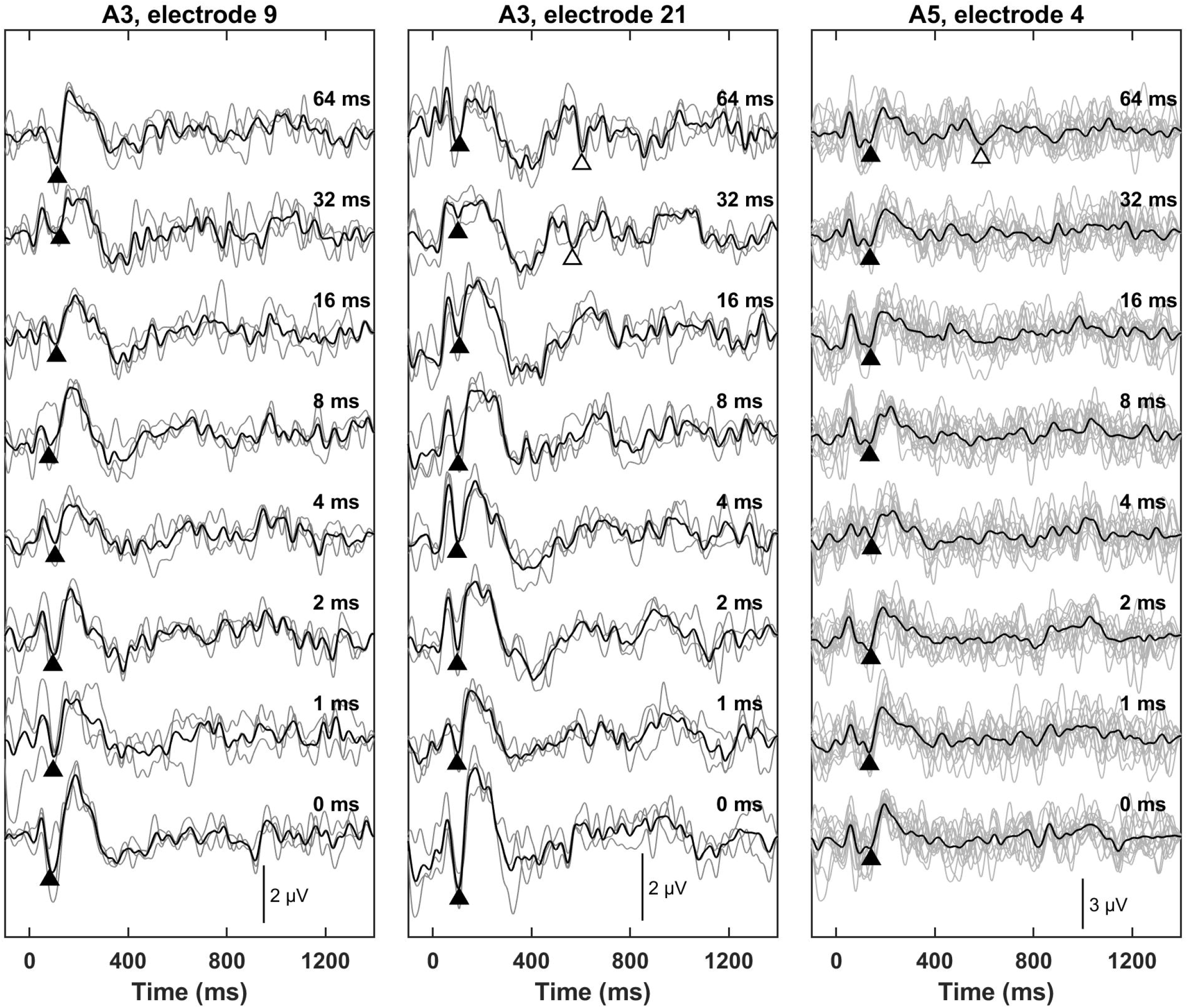
eERPs recorded at three electrode locations in two participants. Each panel shows the results measured at one electrode location. Electrode and participant number were listed at the top of each panel. For each panel, grey lines indicated individual replications recorded in each stimulation condition. Black lines indicated the averaged responses of all replications recorded in the same stimulation condition. Filled and open triangles indicated the N1 peaks of the onset and the eACC identified for each averaged response, respectively. The gap duration tested in each stimulation condition was labeled for these traces.

### GDTs measured for the electrode pairs with different τ_3_s

Figure 4 shows GDTs measured using psychophysical [panel (a)] and electrophysiological [panel (b)] procedures for the electrode pair with different τ_3_ values. Based on our hypothesis, it was expected that higher/larger GDTs would be measured at the electrodes with prolonged neural adaptation recovery (i.e., larger τ_3_s). However, careful inspections of both panels indicated that our results were not always consistent with this expectation. Specifically, while GDTs measured in four ears (A2L, A6R, A8L and A11L) followed this expectation, results measured in another four ears (A1L, A3, A4L and A4R) were opposite to this expected result. In addition, the inconsistency in the results measured using psychophysical and electrophysiological procedures was observed in six ears (A2R, A5R, A7R, A9L, A10L and A10R). Consistent with these observations, the results of related-samples Wilcoxon Signed rank test showed that there was no statistically significant difference in psychophysical (Z = -0.089, p = .929) or objective (Z = -0.865, p = .387) GDTs between the two electrodes with different τ_3_s.

**Figure 4.**
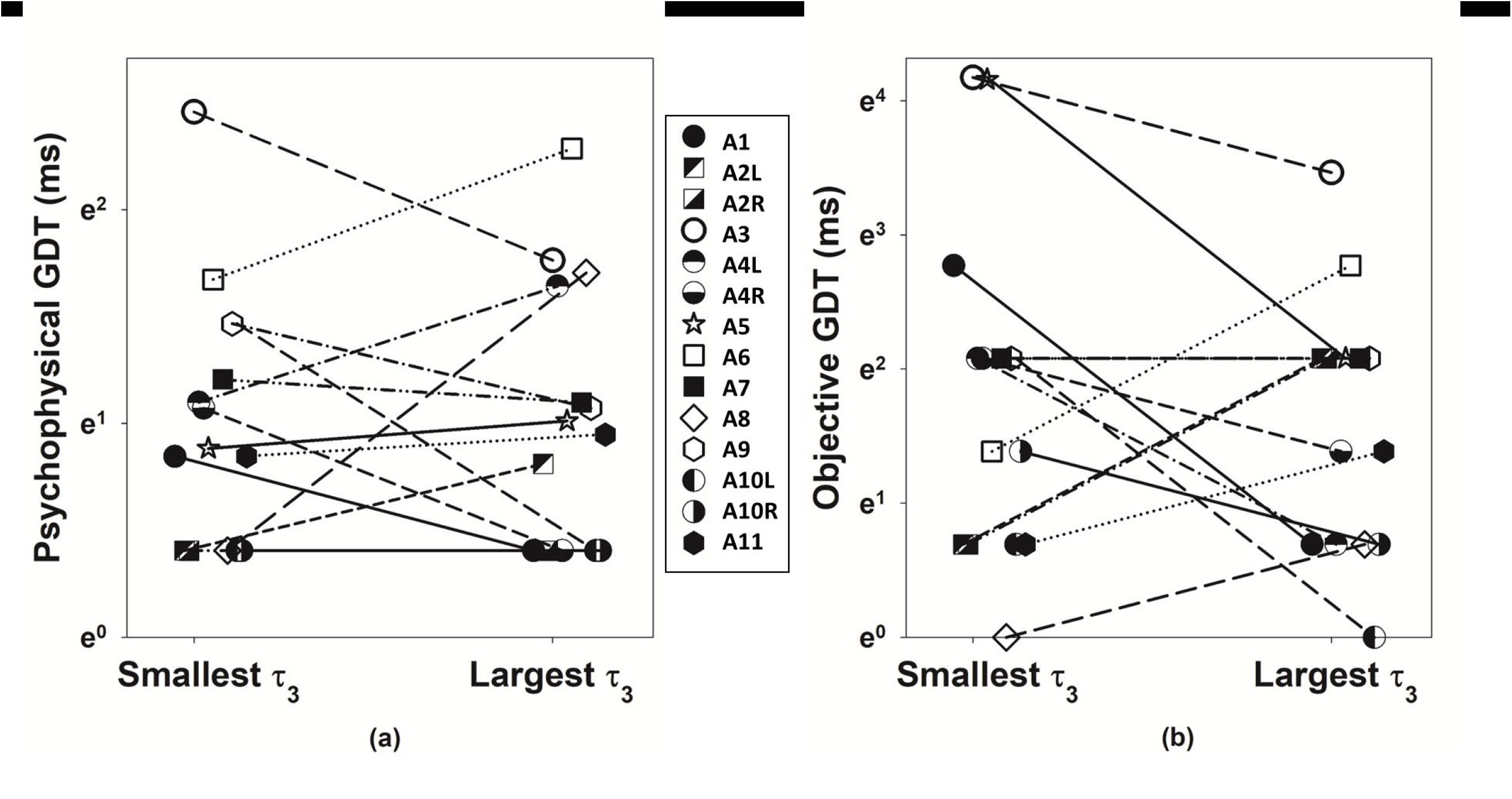
Psychophysical (left panel) and objective (right panel) GDTs measured at the electrode locations with the largest τ_3_ and the smallest τ_3_ in 14 test ears. Each symbol indicates the results measured for one test ear. Results measured in the same test ear are linked by lines.

### GDTs measured for the electrode pairs with different ARRs

Figure 5 shows GDTs measured using psychophysical [panel (a)] and electrophysiological [panel (b)] procedures for the electrode pair with different ARRs. Based on our hypothesis, electrodes with smaller ARRs (i.e., less recovery from neural adaptation) were expected to show larger/higher GDTs. This expected result was not observed in all participants. While data collected in five ears (A2L, A3L, A6R, A8L, and A65L) followed this expected trend, results measured in another two ears (A1L and A4R) demonstrate an opposite trend to the expected result. Similar with the data shown in Figure 4, the inconsistency in the results measured using psychophysical and electrophysiological procedures was observed in seven ears (A2R, A4L, A5R, A7R, A9L, A10L and A10R). The results of related-samples Wilcoxon Signed rank test showed that there was no statistically significant difference in psychophysical (Z = 1.067, p = .286) or objective (Z = 0.393, p = .694) GDTs between the two electrodes with different ARRs, which aligned with the results of visual inspections.

**Figure 5.**
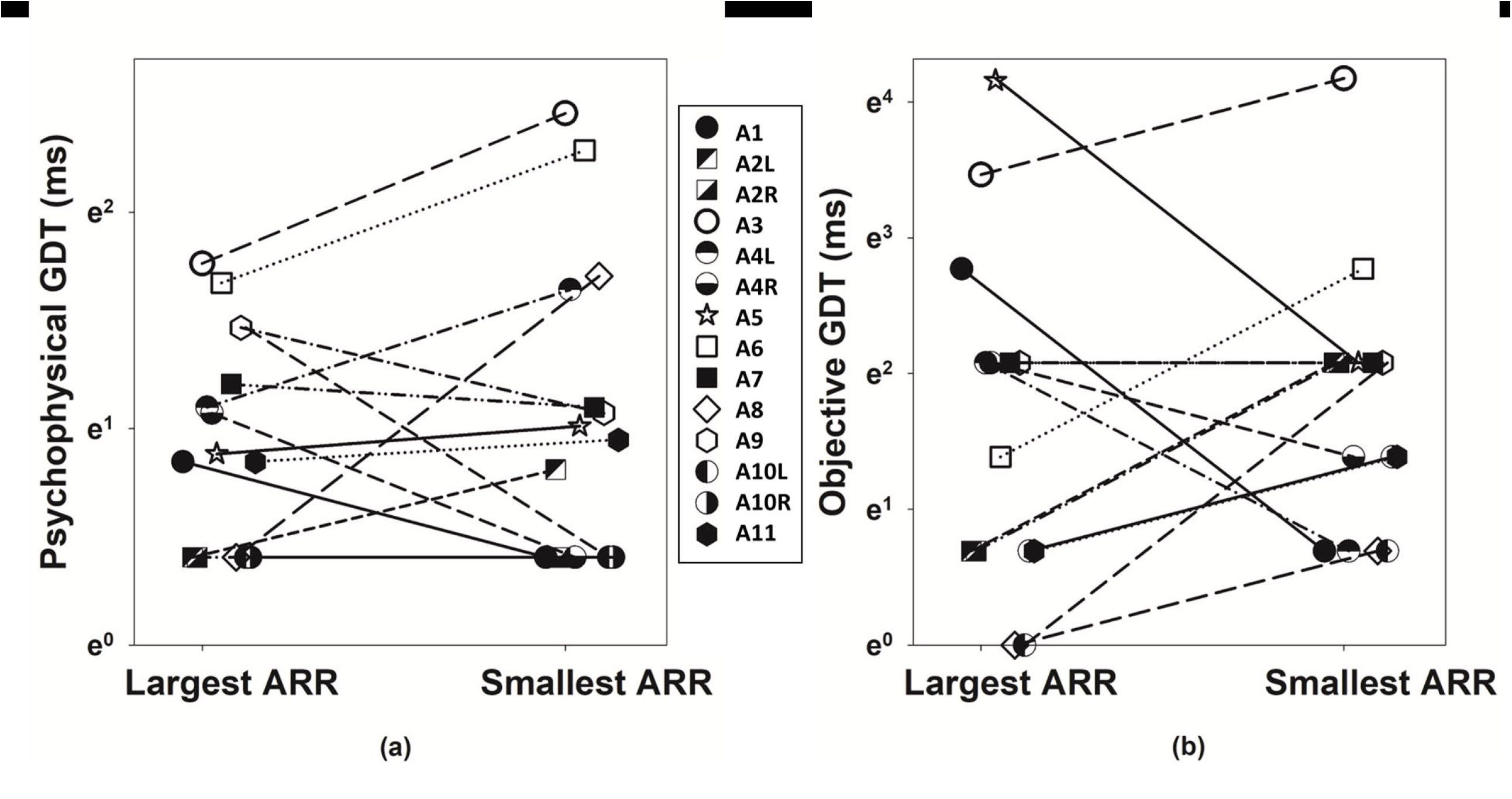
Psychophysical (left panel) and objective (right panel) GDTs measured at two electrode locations with different ARRs in 14 test ears. Each symbol indicates the results measured for one test ear. Results measured in the same test ear are linked by lines.

## DISCUSSION

This study assessed the relationship between neural adaptation recovery of the AN and GDTs measured using psychophysical or electrophysiological procedures in postlingually deafened adult CI users. We hypothesized that the difference in neural adaptation recovery of the AN accounted for the variations in GDTs measured at different stimulation sites within individual CI users. Based on this hypothesis, electrodes with less and/or prolonged recovery from neural adaptation of the AN were expected to show larger GDTs. These expected results were only observed in data collected in some but not all test ears. Overall, results of this study did not support our working hypothesis.

### Psychophysical vs objective GDTs

GDTs measured using psychophysical procedures in this study are consistent with those reported in other studies that used similar testing paradigms and conditions (e.g., Busby & Clark, 1999; Garadat & Pfingst, 2011; Shader et al., 2020). Comparing GDTs measured using electrophysiological procedures across studies is challenging due to the difference in gap durations tested in different studies. Our results showed a significant correlation between GDTs measured using psychophysical and electrophysiological procedures, and a significant difference in GDTs measured using these two procedures with larger GDTs measured using the electrophysiological procedure. Overall, these results are consistent with those reported for acoustical hearing (He et al., 2012). These data generally support the use of electrophysiological measures of the eACC to assess temporal resolution acuity in implanted patients who cannot provide reliable behavioral responses.

Despite these statistically significant results, a large discrepancy between GDTs measured using psychophysical and electrophysiological procedures was observed at three electrode locations (Figures 2 and 3). This discrepancy has been observed in children with auditory neuropathy spectrum disorder (He et al., 2015) and implanted children with cochlear nerve deficiency (He et al., 2018a). In all reported cases, onset ERP responses evoked by acoustical or electrical stimulation could be easily identified. Therefore, the lack of (e)ACC responses was not due to an inadequate stimulation level. Based on the results of some animal studies, it has been proposed that within-channel gap detection is primarily associated with the function of the subcortical auditory structures (Rybalko et al., 2010; Kirby & Middlebrooks, 2012). Consistent with this proposed theory, high electrode impedance and/or poor responsiveness of the AN to electrical stimulation were observed at these three electrode locations tested in this study. However, these limited data are not sufficient to provide any conclusive evidence to support this proposed theory. As a result, the neurophysiological mechanism underlying the disassociation between cortical encoding and auditory perception of within-channel temporal gaps remains unclear.

### Adaptation recovery of the AN vs across-electrode variation in GDT

Our results showed that GDTs could not be predicted based on the speed (Figure 4) or the amount (Figure 5) of adaptation recovery of the AN. Therefore, the difference in adaptation recovery of the AN is unlikely to be the neurophysiological mechanism underlying the variations in GDTs measured at different stimulation sites within individual CI users.

In a recent study, the speed of neural adaptation recovery of AN was found to be associated with speech perception performance in postlingually deafened adult CI users (He et al., 2022b). As a measure of temporal processing acuity, within-channel GDT has also been shown to be correlated with speech perception performance in CI users (e.g., Goldsworthy et al., 2013; Blankenship et al., 2016; Xie et al., 2022). It needs to be pointed out that these significant results were either based on the data averaged across multiple CI electrodes or measured using sound field presentation that activated multiple electrodes across the electrode array. In this study, the relationship between adaptation recovery of the AN and GDT was assessed using a within-subject design and was based on results measured at individual CI electrodes. Therefore, the lack of association between adaptation recovery of the AN and within-channel GDT measured at individual CI electrodes in this study does not necessarily contradict these previously published, significant results.

### Potential Study Limitations

This study has two limitations. First, only 11 participants were tested in this study and both ears were tested in three participants with bilateral CIs. This small sample size, along with the potential lack of independence between results measured from the two ears of the same participant, might have affected the results of this study. Second, different stimulation durations were used in different testing paradigms. Specifically, the stimulation duration used for assessing adaptation recovery of the AN was 100 ms. The pulse train durations used in psychophysical measures of GDT and eERP recordings were 500 ms and 800 ms, respectively. In this study, eCAP results were used to select the electrode pair for measuring GDTs instead of directly comparing with GDT data. In addition, a duration difference between 500 ms and 800 ms should not affect GDT results when all other stimulation parameters are the same (Busby & Clark, 1999). Therefore, using different stimulation durations in different tests is unlikely to affect the results of this study.

## CONCLUSIONS

Electrophysiological measures of the eACC can potentially be used to assess within-channel GDTs in implanted patients who cannot provide reliable behavioral responses. Neural adaptation recovery of the AN is not a crucial factor for determining within channel GDT in postlingually deafened adult CI users. Further studies are warranted to determine whether variations in GDTs measured at different stimulation sites within individual CI users are due to factors that are central to the AN within the auditory system.

## Data Availability

Please contact the corresponding author to discuss access to the data presented in this study.

## ACKNOWLEDGMENTS

This work was supported by an R01 grant from NIDCD and NIGMS (1R01DC016038). We gratefully thank all study participants for participating in this study.

